# Abnormal visual-vestibular cerebro-cerebellar connectivity in persistent postural-perceptual dizziness

**DOI:** 10.64898/2026.07.31.26359398

**Authors:** Hannah Schewe, Renana Storm, Hannah Keller, Antonia Frings, Pia Herborn, Viktoria Wrobel, Skadi Gerkensmeier, Christoph Helmchen, Andreas Sprenger

**Author notes:** Hannah Schewe and Renana Storm shared first authorship. **Corresponding author:** Christoph Helmchen, MD, Department of Neurology; University of Lübeck; University Hospital Schleswig-Holstein; Campus Lübeck; Ratzeburger Allee 160; D-23538 Lübeck.

## Abstract

**Objective:** Patients with persistent postural-perceptual dizziness (PPPD) perceive unsteadiness associated with abnormal ego- or visual motion perception. Maladaptive multisensory integration and visual dependency have been proposed implicating abnormal functional connectivity (FC). Based on the crucial impact of the cerebellum for prediction errors of sensory perception, we specifically investigated group-related differences in FC between visual cortex, multisensory vestibular cortical areas and the cerebellum.

**Methods:** Resting-state activity (seed-based functional connectivity using a priori regions of interest and fractional amplitude of low-frequency fluctuations, fALFF) was compared between a large cohort of 53 PPPD patients and 54 age- and sex-matched HC and related to various disease parameters and baseline postural sway speed.

**Results:** FC between the patients’ posterior insula and the visual (lingual) cortex, as well as the supramarginal gyrus was lower. In contrast, it was markedly larger between the patients’ (i) visual motion-related area MT/V5 and cerebellar Crus I, (ii) multisensory operculum and caudal vermis, (iii) OP3 and Crus II, and (iv) inferior parietal lobe and vermis. fALFF was larger in the patients’ right hippocampus.

**Interpretation:** The abnormally low cortical visual-vestibular FC may provide a neural substrate for the patients’ maladaptive multisensory integration with aberrant inter-sensory reweighting. The larger cerebro-cerebellar FC with cerebellar areas involved in sensory and postural prediction errors control (required for updating internal models of motion) might account for the abnormally high sensory precision weighting leading to the patients’ altered egomotion perception. Longitudinal studies are required to investigate whether the larger cerebro-cerebellar connectivity is compensatory or dysfunctional in nature.

## Introduction

Persistent postural-perceptual dizziness (PPPD) is a common chronic neuro-otologic disorder classified by the Bárány society^1^. PPPD is often preceded by vestibular disorders. Symptoms can be exacerbated by upright posture, active or passive movement, and exposure to moving or complex visual stimuli. According to current pathophysiological models^2, 3^, patients experience a chronic misperception of subjective and objective stability. This is thought to arise from abnormal egomotion processing associated with excessive body vigilance and altered visual and vestibular (i.e., egomotion) perception thresholds^4–7^. Therefore, abnormal visual-vestibular integration could be a feasible mechanism underlying PPPD and may indicate altered functional connectivity (FC)^8^.

Previous resting-state functional MRI studies on PPPD revealed inconsistent results which may be related to the sample size^9^ and/or the various resting-state analysis techniques applied^7–15^. For example, FC between the occipital pole visual network and sensorimotor or spatial networks has been reported to be increased^9, 16^ and decreased^8^, with both findings being linked to abnormal visual dependence and impaired multisensory integration, key neural mechanism proposed for PPPD.

The cerebellum is crucially involved in human egomotion perception and postural control^17^. Particularly, the caudal vermis (uvula) integrates self-motion signals with its strong functional connections to the cingulate sulcus visual area (CSv), the posterior cingulate area (pCi), the precuneate motion area (PcM), as well as the temporo-parietal cortex including the visual posterior sylvian area (VPS) and the supramarginal gyrus (SMG). These projections are likely to form feedback loops that continuously update visual-spatial and vestibular information, thereby supporting self-referential spatial awareness. In particular, Crus I has been implicated in predicting the sensory consequences of head and body movements and in comparing predicted with incoming sensory feedback^18–20^. Likewise, the posterior cerebellum provides temporal signals to parietal networks involved in spatial orienting^20^.

In this predictive framework, expected and actual sensory signals are continuously compared, with matches leading to attenuation of predictable input and mismatches generating prediction errors that update internal models of motion. Given its central role in these computations, altered cerebellar processing could thereby influence sensorimotor networks underlying postural control^20^. Abnormal amplification or a failure in the attenuation of sensory prediction errors have been suspected in PPPD^7, 21^.

Accordingly, we hypothesized that abnormal FC between cortical visual-vestibular areas is associated with altered cerebro-cerebellar connectivity, specifically the caudal vermis and cerebellar Crus I and II being involved in error prediction. Abnormal visual-vestibular integration has already been speculated to be related to a modified cerebellar network connectivity in PPPD precursor entities^14^. Recently, some lines of evidence for altered cerebellar FC have been provided with inconsistent results, i.e., decreased^8^ and increased^13^ FC between visual and opercular networks and the cerebellum. The functional meaning remained unclear.

## Methods

### Participants

The final sample comprised 53 patients (female: 34, male: 19, meanage= 45.5 years, SDage = 11.6 years) and 54 age- and sex-matched healthy controls (HC; female: 33, male: 21, meanage = 44.3 years, SDage = 12.8 years). Patients were recruited at the University Centre for Dizziness and Vertigo in Lübeck and met the Bárány Society diagnostic criteria for PPPD^1^. Exclusion criteria included cerebral lesions, substance abuse, severe psychiatric disorders, visual impairment, and current comorbidities including migraine, vestibulopathy, benign paroxysmal postural vertigo, tinnitus, or acute hearing loss. All participants had completed at least nine years of formal education. MRI datasets were excluded if head motion exceeded 3 mm of translation or 3° of rotation, if the mean voxel displacement exceeded 1.5 mm, or if more than 50% of volumes exceeded a framewise threshold of 0.6 mm. Overall, 4 participants were excluded prior to the final analyses because of clinical or technical exclusion criteria.

The study protocol was conducted in accordance with the Declaration of Helsinki and approved by the local ethics committee of the University of Lübeck (AZ 17–036, AZ 21–098). Written informed consent was obtained from all participants.

### Experimental procedure

Participants were asked to complete the Niigata PPPD Questionnaire^22^ (NPQ), the Athens-Lübeck-Questionnaire^23^ (ALQ) for PPPD-subtyping, the Hospital Anxiety and Depression Scale^24, 25^ (HADS), the NEO-Five-Factor Inventory^26^ (NEO-FFI), the short form of the Motion Sickness Susceptibility Questionnaire^27^ (MSSQ), and the Edinburgh Handiness Questionnaire^28^ (EHQ). 95% of all participants were right-handed. All questionnaires were completed at home prior to their MRI measurement.

Prior to the study, participants underwent a thorough bedside neurological examination as well as the following tests: video-based quantitative head impulse test using the EyeSeeCam® HIT System (Autronics, Hamburg, Germany), subjective visual vertical (normal reference: ± 2.5°), and ocular-vestibular evoked myogenic potentials (for further details see^5, 29, 30^). 14 patients had a previous vestibular episode of peripheral paroxysmal positional vertigo or vestibular neuritis, but no participant showed abnormal values or clinical signs at the time of MRI recordings.

### Image acquisition

Structural and functional MRI was performed at the Center of Brain, Behavior and Metabolism (CBBM) Core Facility for Magnetic Resonance Imaging using a 3 T Siemens Magnetom Skyra scanner equipped with a 64-channel head-coil. Functional images were acquired applying a single-shot gradient-recalled echo-planar imaging (GRE-EPI) sequence sensitive to BOLD contrast (TR = 1020 ms; TE = 29 ms; flip angle = 70°; in-plane resolution 3 × 3 mm²; 192 × 192 mm² field of view; 60 transversal slices; 3 mm slice thickness; simultaneous multi-slice factor 4); 465 volumes were recorded. Additionally, structural images of the whole brain using a 3D T1-weighted MP-RAGE sequence were acquired (TR = 1900 ms; TE = 2.44 ms; TI = 900 ms; flip angle 9°; 1 × 1 × 1 mm³ resolution; 192 × 256 × 256 mm³ field of view; acquisition time 4.5 min). T1-weighted images were inspected for pathological findings by a neuroradiologist. Head movements were minimized using ear pads (Multipad Ear, Pearltec Technology AG, Schlieren [CH]). Ear plugs were used for noise cancellation. Eye movements were recorded via a MR-compatible video based eyetracker (Eyelink 1000 Plus, 1000 Hz sampling rate, SR Research Ltd., Ottawa, ON/Ca). Participants were positioned in a standard procedure with a specially designed eye-tracking mirror placed above their face so they could see the monitor behind them. For resting state image acquisition, a central orange dot was presented on the monitor (NordicNeuroLab LCD Monitor, 32-inch diameter, screen resolution: 1920 × 1080 pixel, 60 Hz refresh rate). Participants were asked to fixate the dot and clear their minds of any thoughts. Eye movements were used to monitor participants’ vigilance and fixation performance.

### Preprocessing

Preprocessing and first level analysis were performed using *HALFpipe 1.3.2*^31^. The first 5 volumes were removed from functional images. The preprocessing steps included skull stripping, slice timing and susceptibility distortion correction using field maps, co-registration, and spatial normalization to the MNI152NLin2009cAsym template (2 × 2 × 2 mm³), which is defined as standard space in HALFpipe.

For the seed-based FC analysis, the images were further denoised using spatial smoothing (6 mm full width half maximum Gaussian kernel), grand mean scaling (10,000) and a Gaussian-weighted temporal filtering (high-pass filter width 125 s). Each seed had to be covered by at least 80% brain voxels in order to be included in subsequent analysis. To reduce motion artefacts and physiological noise, the time series of six motion parameters, white matter, and cerebrospinal fluid were regressed out.

For fractional amplitude of low-frequency fluctuation analysis (fALFF), we used spatial smoothing (6 mm full width half maximum Gaussian kernel), grand mean scaling (10,000), and frequency based temporal filtering (low-pass: 0.01 Hz; high-pass: 0.1 Hz). For denoising, we regressed the time series of six motion parameters, motions outliers (motion scrubbing; threshold: framewise displacement = 0.5 mm), white matter, and cerebrospinal fluid.

Following preprocessing and first-level analyses, data quality was assessed using the data quality assessment provided by HALFpipe^31^. Each subject was reviewed by at least two researchers.

### Seed-based functional connectivity analysis

Seed-based FC analyses were performed using a seed-to-voxel approach. Regions of interest (ROI) were defined a priori based on previous literature and task-based analyses of the same dataset^17, 21^. Specifically, ROIs were selected from cortical regions associated with the visual and vestibular networks as well as the Default Mode Network (DMN). ROI masks were derived from the Julich Brain Atlas^32^ and the Harvard-Oxford Atlas^33^. All ROIs were defined in MNI152NLin2009cAsym space. A complete list of ROIs is provided in Supplementary Table S1. The average BOLD time series of the ROI is used as a regressor in a first-level GLM for each voxel of the brain^31^. We obtained whole-brain FC maps for each ROI per subject, which revealed the strength of the connectivity between the ROI and every voxel in the brain^31^.

### Fractional amplitude of low frequency fluctuations (fALFF) analysis

We performed a voxel-wise fALFF analysis^34^ as a measure of spontaneous activity. Accordingly, the variance of the amplitude in the low-frequency range of the BOLD signal is calculated by dividing the power in the low-frequency range (0.01-0.1 Hz) by the power across the entire frequency range. Subject-specific fALFF maps are z-scaled by default in HALFpipe^31^.

### Posturographic measurements

We recorded participants’ posturographic sway parameters outside the scanner: Participants stood on a Kistler force platform (Model 9260AA6, Kistler Instrumente AG, Winterhur Switzerland; 50 cm width, 60 cm length) equipped with piezo-electric 3-component force sensors for recording postural changes. We obtained the postural sway speed on a firm platform with (fixation gaze straight ahead) and without (eyes closed) visual control as a parameter for postural control. Results are presented as the median postural sway speed (mm/s), calculated from the vector sum center-of-pressure (CoP) displacement in the mediolateral and the anteroposterior directions using Matlab® (R2024b, The Mathworks, Natick/MA). After each posturographic measurement, participants were asked to rate their perceived sway on a visual analogue scale from 0 % (*did not feel anything*) to 100 % (*almost fell off the platform*). Part of the posturographic data during various experimental conditions have been published elsewhere^30^.

### Statistical analysis

All statistical analyses were performed in custom Python scripts using nilearn^35^ for fMRI group comparisons (*Python 3.12, nilearn 0.13.1; scipy 1.15.3*). We compared questionnaire scores and behavioral parameters between HC and patients by using Mann-Whithey-*U*-Tests for non-normally distributed data or Welch’s *t*-test for normally distributed data. To compare fMRI results between groups, we used two-sided independent *t*-tests with non-parametric cluster-based permutation inference^36^, which provides well-controlled false-positive rates^37^. The cluster-forming threshold was set at *p* < 0.001. Statistical significance was assessed using a cluster-mass permutation test with family-wise error rate (FWER) correction (*p* < 0.05, 10,000 permutations per test). We analyzed only voxels for which at least 80 % of participants provided valid data. To interpret results, corrected −log10(*p*) maps were multiplied by the sign of the corresponding *t*-statistic, resulting in signed, corrected significance maps that preserved the direction of group differences. Reported anatomical labels were assigned based on the nearest automated anatomic labeling (AAL) atlas region to the peak voxel coordinate^38^. For findings in the cerebellum, reported anatomical labels were derived using the spatially unbiased atlas template of the cerebellum and brainstem^39, 40^ (SUIT atlas), based on the region that showed the largest overlap with each significant cluster.

For correlation analyses between clusters showing altered FC or fALFF and questionnaire scores or disease-related parameters, non-parametric Spearman’s rank correlations were calculated. To account for multiple comparisons, p-values were Bonferroni-corrected for the number of included brain clusters. Pairwise correlations among all clusters with altered FC or fALFF in patients were assessed using Spearman’s rho and Bejamini-Hochberg false discovery rate (FDR) procedure at *q* < 0.05, based on the total number of pairwise correlations.

Analysis scripts were initially created using GPT-5.5 (accessed June 2026). All AI-generated code was carefully reviewed, tested, and revised by the authors, who confirmed the accuracy and reproducibility of the results.

## Results

### Disease-related scores and personality traits

Patients had an average disease duration of 32 months (SD = 33.1 months). All results from the questionnaire data for HC and patients are presented in Supplementary Table S2. Patients showed significantly larger NPQ (*z* = −8.65, *p* < 0.001) and ALQ (*z* = −8.79, *p* < 0.001) scores, HADS anxiety and depression scales (HADS-A: *z* = −6.45, *p* < 0.001; HADS-D: *z* = −6.66, *p* < 0.001). Patients showed larger values on the subscales of NEO-FFI for neuroticism (NEO-N: *t*(103.18) = −5.56, *p* < 0.001), but lower values for extraversion (NEO-E: *t*(104.97) = 3.47, *p* < 0.001) and agreeableness (NEO-A: *t*(104.56) = −2.0, *p* = 0.048). No group differences occurred in the MSSQ, NEO-FFI for openness (NEO-O) and conscientiousness (NEO-C). Patients differed in terms of the speed of postural sway with their eyes open (*z* = −2.07, *p* = 0.039) and with their eyes closed (*z* = −3.09, *p* = 0.002). In both conditions, they also perceived their sway as more pronounced and therefore exhibited elevated subjective sway ratings (eyes open: *z* = −3.69, *p* < 0.001; eyes closed: *z* = −3.43, *p* < 0.001).

### Altered functional connectivity in PPPD

Seed-based FC analyses revealed several significant clusters for the group comparison (Supplementary Table S3). There were a number of significant clusters with stronger FC in patients, largely with the cerebellum: between left V5 (seed) and right Crus I (*p* = 0.018) as well as between right V5 (seed) and left Crus I (*p* = 0.040; **Fig 1A**). Connectivity between left inferior parietal lobe (IPL) PFcm (seed) and the vermis (*p* = 0.008) as well as to a subcortical cluster in right putamen (*p* = 0.029; **Fig 1B**) was larger in patients. Right IPL PFcm (seed) showed larger connectivity with a cluster in the right caudate (*p* = 0.033). Additionally, two further clusters with stronger connectivity between the parietal operculum (OP1, seed) and cerebellar vermis were identified: left OP1 (*p* = 0.009) and right OP1 (*p* = 0.017; **Fig 1B**). FC between right OP3 (seed) and left Crus II (*p* = 0.049) as well as between left OP4 (seed) and the right thalamus (*p* = 0.012; **Fig 1C**) was also larger in patients. In contrast, patients showed reduced FC between the left posterior insula (Ig 2, seed) and the left lingual gyrus (LingG, *p* = 0.022) as well as the left supramarginal gyrus (SMG, *p* = 0.046; **Fig 1D**).

**Figure 1.**
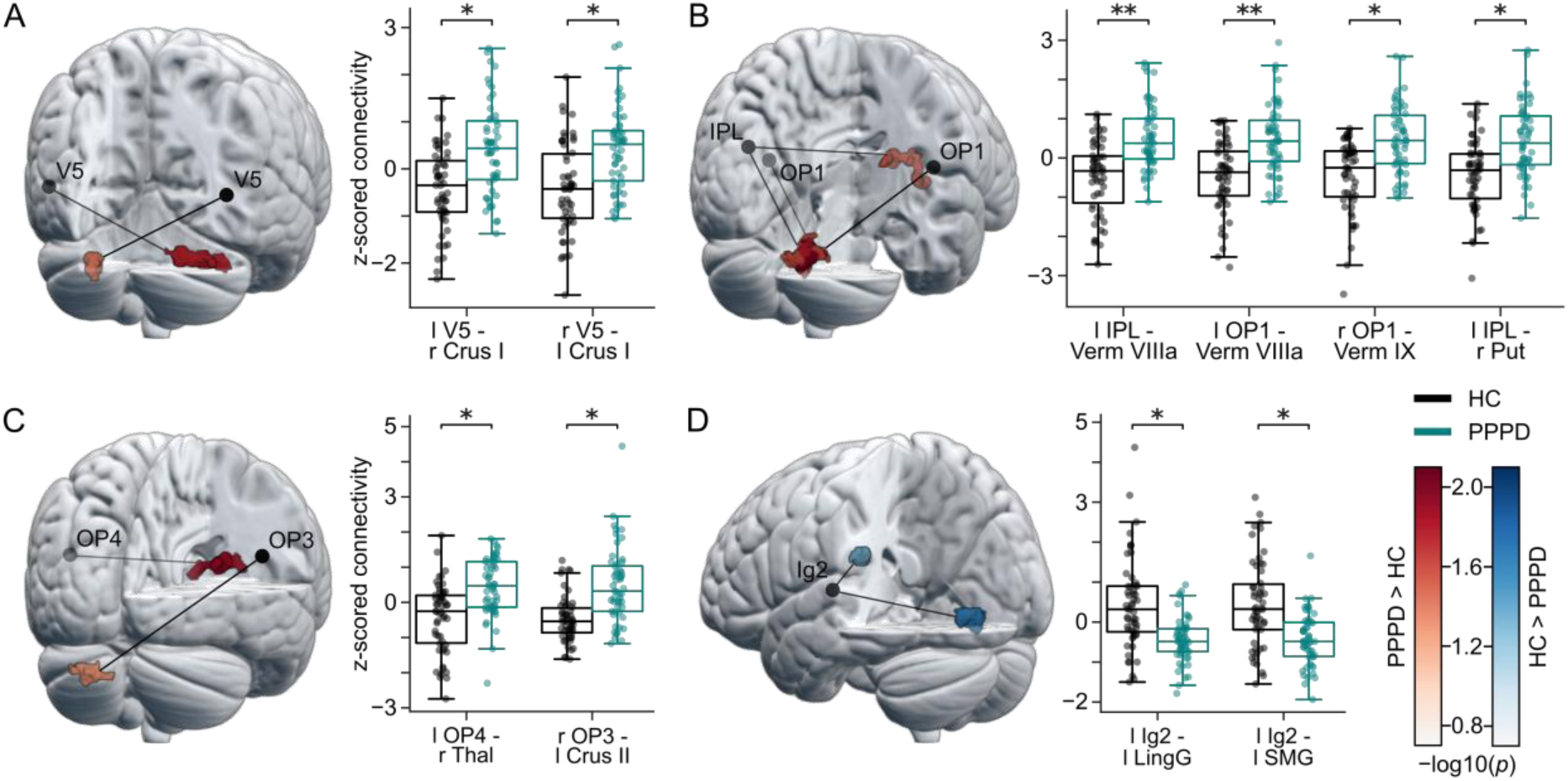
Altered clusters of seed-based functional connectivity in PPPD. **a)** Left: Whole-brain rendering showing clusters of significant group differences between PPPD patients and HC. Red clusters indicate larger FC in patients, with the color gradient representing significance level (−log10(*p*)). Black dots represent the ROIs, and solid black lines connect each ROI to its corresponding significant cluster. Right: Boxplots showing the median z-scored FC values within the significant clusters for each group. Dots show median FC for single subjects. Asterisks indicate significant differences based on non-parametric cluster-based permutation tests (* *p* < 0.05, ** *p* < 0.01, *** *p* < 0.001). HC are shown in black and patients are shown in green. Panels **b**, **c** and **d** follow the same layout as panel **a**. **d)** Blue clusters indicate lower FC in PPPD patients, with the color gradient representing significance level (−log10(*p*)). Abbreviations are listed at the end of the manuscript.

**Fig 2** provides an enlarged anatomical visualization of the findings within the cerebellum. Specifically, **Fig 2A** shows axial cerebellar slices with all significant vermal clusters (seeds: left IPL PFcm, left and right OP1), whereas **Fig 2B** displays the corresponding cerebellar surface map with all significant cerebellar clusters. The percentage overlap of each cluster with SUIT atlas regions is reported in Supplementary Table S4. Finally, **Fig 3** presents an overview of all altered seed-to-cluster connectivity.

**Figure 2.**
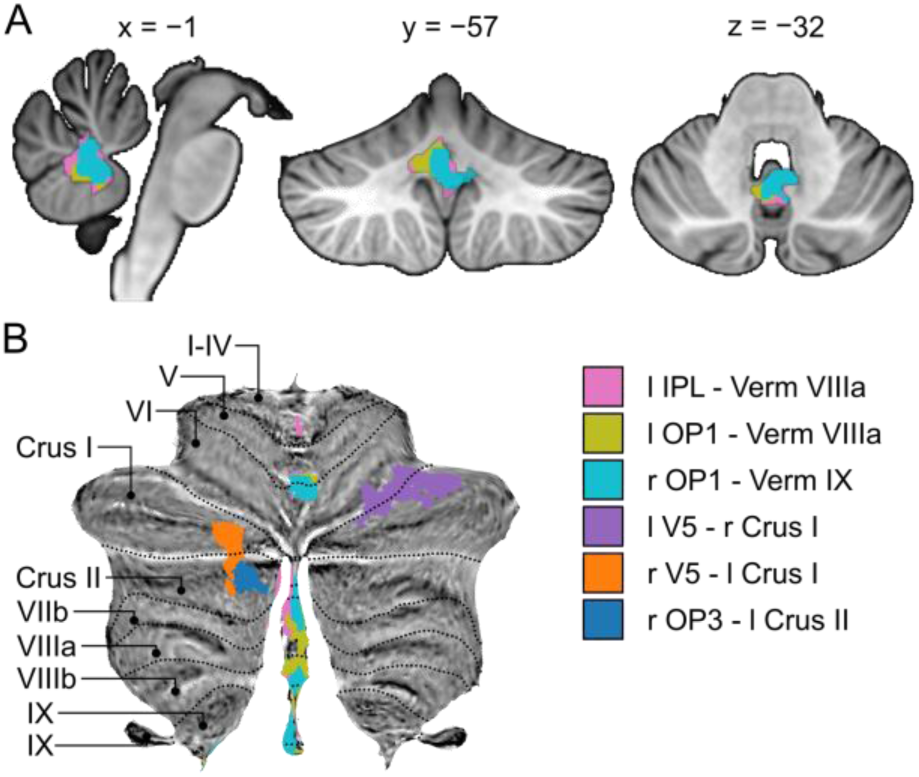
Cerebellar clusters of significant group differences in seed-based functional connectivity. All clusters shown represent larger FC in PPPD patients compared to HC. The same legend applies to panels **a** and **b**. For each legend entry, the first region listed represents the seed region (not shown), whereas the second represents the corresponding significant cluster, identified according to the cerebellar region with the largest overlap in the SUIT atlas. **a)** Sagittal (x = −1), coronal (y = −57), and axial (z = −32) views of the significant clusters located in the vermis. **b)** Flatmap of the SUIT atlas showing the significant cerebellar clusters located in the vermis, Crus I and Crus II.

**Figure 3.**
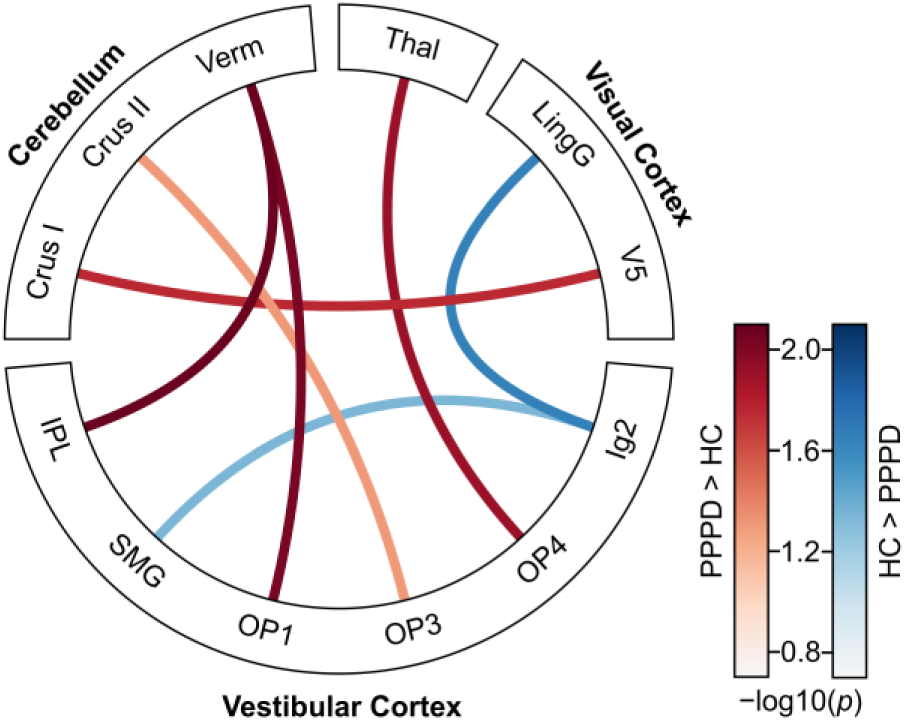
Summary of findings for group differences in seed-based functional connectivity. Red lines indicate larger and blue lines indicate lower FC in PPPD patients relative to HC, with the color gradient representing significance level (−log10(*p*)). Lateralization was omitted in this schematic representation for clarity.

### Stronger resting-state activity in the hippocampus in PPPD

Using whole brain voxel-wise fALFF analysis, group comparison revealed one cluster of significantly larger resting-state activity in the right hippocampus in patients (*p* = 0.048; **Fig 4**).

**Figure 4.**
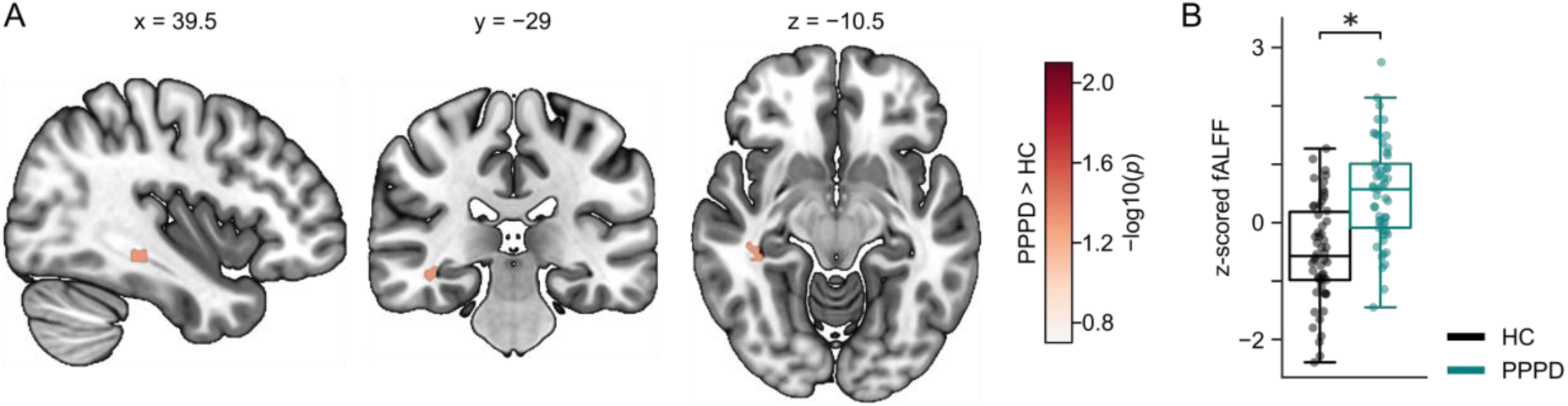
Significant cluster of group difference in fALFF analysis. **a)** Sagittal (x = 39.5), coronal (y = −29), and axial (z = −10.5) views of the significant cluster located in the right Hippocampus. Red cluster indicates larger fALFF value in patients, with the color gradient representing significance level (−log10(*p*)). **b)** Boxplot showing the median z-scored fALFF value within the significant cluster for each group. Dots show median fALFF for single subjects. Asterisk indicates significant difference based on non-parametric cluster-based permutation test (* *p* < 0.05, ** *p* < 0.01, *** *p* < 0.001). HC are shown in black and patients are shown in green.

### Correlation analysis of functional connectivity and clinical data

FC between right OP3 and Crus I increased with age of the PPPD patients (*ρ* = 0.39, *p* = 0.037). There was no correlation in HC (*ρ* = −0.06, *p* = 1.0). No other correlations between FC or fALFF values and demographic data (age, disease duration), data from clinical questionnaires (ALQ, NPQ, HADS, MSSQ), or behavioral measures (postural sway speed, subjective sway ratings) remained significant after Bonferroni-correction.

Pairwise correlation analyses of FC and fALFF values extracted from clusters showing significant group differences revealed several positive correlations after FDR correction. Specifically, the left OP1-vermis FC increased with larger FC values for the right OP1-vermis (*ρ* = 0.80, *p* < 0.001) and the IPL-vermis FC (*ρ* = 0.80, *p* < 0.001). The right OP1-vermis FC increased with the IPL-vermis FC (*ρ* = 0.70, *p* < 0.001). Furthermore, FC between V5 and contralateral cerebellar Crus I on both sides correlated with each other: FC between left V5 and right Crus I became larger with larger values for the FC between right V5 and left Crus I (*ρ* = 0.54, *p* < 0.001). In addition, FC between right V5 and left Crus I was positively correlated with fALFF in the right hippocampus (*ρ* = 0.37, *p* = 0.046). All pairwise correlations are shown in Supplementary Figure S1.

## Discussion

Our main findings revealed reduced FC between the patients’ multisensory vestibular cortical areas (posterior insula and SMG) and visual cortex (LingG) as well as larger FC between the patients’ visual motion area MT/V5, multisensory operculum, and IPL, respectively, and the cerebellum. Specifically, the identified cerebellar clusters were located in regions linked to egomotion perception, visuomotor and postural control, as well as predictive processing, including sensory prediction and prediction error signaling. Correlations between the down-(cortical visual-vestibular) and upregulated (cerebro-cerebellar) FC did not confirm our initial hypothesis that both FC changes are functionally interdependent.

### Reduced visual-vestibular seed-based functional connectivity in PPPD

Reduced FC in the multisensory operculum has previously been reported in PPPD precursor syndromes^16, 41^. In PPPD, however, studies have reported inconsistent findings regarding FC between multisensory operculum and visual cortex, with both larger^9, 16, 41^ and lower FC^8, 12^. We specifically identified a reduced FC between the patients’ lingual gyrus in the visual cortex and the posterior insular cortex. This is in line with related studies using seed-based FC analysis^12^ and graph theoretical analysis^8^. Yagi et al. showed reduced FC between the posterior insula, formerly called the parieto-insular vestibular cortex in the monkey^42, 43^, and the lingual gyrus. Li et al. used defined networks and showed a lower interaction between opercular and visual networks. Reduced seed-based FC was also reported between precuneus and the visual cortex and corpus callosum^9^. Both, lingual gyrus and precuneus, provide basic visual information of target location and content to the dorsal and ventral stream required for spatial orientation^44^. In contrast to Li et al.^9^, we did not identify altered connectivity using precuneus as a seed. In sum, functional and network connectivity analyses in various studies indicate abnormal cortical visual-vestibular connectivity in PPPD.

Dysfunctional visual-vestibular FC with the medial occipito-temporal (lingual) gyrus in our data could impair basic visual processing, motion discrimination, visual memory, visual attention^45^, and imagery^46^. That may account for elevated visual motion detection thresholds^5^ and visual dependency in PPPD, i.e., the increase of patients’ dizziness on exposure to complex visual stimuli^1^. Pathophysiologically, this may lead to visual-vestibular incongruencies by disrupting the spatial tuning, reference frames, and temporal dynamics that normally support visual-vestibular integration^47^.

Spatial orientation and egomotion perception require an intact visual-vestibular interaction in order to distinguish egomotion from motion of the surrounding environment^48^. One mechanism underlying this distinction is the so-called reciprocal cortical visual-vestibular inhibitory interaction, which deactivates the visual cortex during vestibular stimulation and vice versa^49^. Following a prolonged vestibular sensation, such as acute vestibulopathy, multisensory inputs are dynamically reweighted to maintain postural control and egomotion perception^50^. The attenuated vestibular input is compensated by an increased visual and somatosensory dependance for postural control. This multisensory reweighting process remains highly adaptive, whereas reduced visual-vestibular FC may delay the down-weighting of visual input during postural control. This could contribute to abnormal egomotion, as the ability to distinguish self-from object-motion or from motion in the visual environment is reduced^5^. Patients’ visual dependence of dizziness is strongly influenced by posture^51^, the type of sensory stimulation (visual versus multisensory), and attentional allocation^52, 53^. In this respect, it is noteworthy that the reduced visual-vestibular FC was limited to vestibular processing areas in Ig2 in the posterior insula and the SMG, but did not affect the core regions of vestibular processing in the parietal operculum (OP1-4)^54^.

Seed-based connectivity in the SMG was also found to be reduced in patients with bilateral^55^ and acute unilateral vestibular failure which partially reversed over a period of three months after improvement^56^. Only four patients of our sample had a history of vestibulopathy, however, no participant showed quantitative signs of vestibular hypofunction at the time of MRI recordings. Abnormally reduced FC in the SMG could persist as a consequence of a previous bilateral vestibulopathy^55^. In PPPD, the SMG not only appears to exhibit reduced FC with the posterior insula (Ig2), but it is also more strongly activated during vestibular stimulation (GVS) compared to HC^21^. Ig2 serves as a hub to integrate nociceptive and somatosensory information^57^. Its interoceptive function is extended by vestibular information through functional connections with the SMG, which were reduced in our PPPD participants. Although this reduced FC could contribute to abnormal egomotion in PPPD, it was not associated with individual egomotion perception ratings. Reduced visual-vestibular connectivity between Ig2 and the lingual gyrus and SMG, may give rise to sensory prediction errors that require continuous updating of internal models. Functionally, this altered cortical visuo-vestibular connectivity may affect visual processing, motion discrimination, visual memory, and visual attention. Notably, the increased FC between core vestibular regions (OP1, OP3, IPL) and the cerebellum may therefore become important, as the cerebellum is involved not only in sensorimotor but also in sensory-perceptual learning and the processing of prediction errors^18–20^.

### Larger cerebro-cerebellar seed-based functional connectivity in PPPD

We identified larger FC between both the visual cortex and the core vestibular processing areas in the patients’ operculum and the cerebellum. This contrasts a functional network analysis^8^, but is in line with a recent seed-based FC study^13^. Li et al. used network-based statistics and graph theoretical analyses to speculate that lower FC between the patients’ visual cortex and the cerebellum may lead to postural instability as it increased with the level of subjective dizziness in daily life (DHI)^8^. However, these were global efficiency network analysis without defined cerebellar clusters or region-specific hypotheses.

Kang and coworkers defined intrinsic connectivity networks (ICN): the visual network (VIS) covered large parts of the entire visual cortex, the multisensory vestibular cortex (MVC) covered posterior perisylvian cortex, OP2, and the whole temporo-parietal junction (TPJ), and the cerebellum network (CB) encompassed the entire cerebellum. Our analysis pinpointed defined areas within the ICN that may account for the altered FC in their analysis.

Notably, patients showed larger FC between OP1 and IPL with the caudal vermis, including uvula (**Fig 2 and 3**). The operculum (OP1, OP2) and the uvula are core structures of the non-human^58^ and human egomotion networks^17, 59^. The vermis is involved not only in postural control^60^ but also in predictive adjustments that compensate for external and internal perturbations^18^. To this end, it generates forward internal models that predict the sensory consequences of actions, thereby compensating for the temporal delays inherent in feedback control of motor commands and perceptual processes^61^. Likewise, posterior and caudal vermal regions encode vestibular information and integrate it with extravestibular signals to generate internal models of eye, head, and body movements, as well as their spatial orientation relative to gravity^62^. Accordingly, cerebellar dysfunction may impair the anticipatory postural adjustments required to compensate internally generated postural sway^63^. Through these internal predictive models, the cerebellum is also involved in learning to anticipate unexpected sensory consequences during voluntary self-motion^64^. PPPD patients have altered visual and vestibular motion perception thresholds and are prone to recognize egomotion even in a non-motion condition^5^. This may reflect prediction errors of forthcoming perceptual events in which the cerebellum is involved^20^. Abnormal dizziness, including perceived unsteadiness, may result from abnormal precision weighting of prediction errors, driven by altered sensory perception thresholds that lead to inaccurate predictions of posture, i.e., postural misperceptions^29, 65^. Based on this, the question arises, whether the larger opercular-vermal FC in PPPD patients is beneficially adaptive or maladaptive in nature.

During visual and vestibular egomotion-compatible stimulation, FC between the operculum and uvula increases^17^. One might speculate that the suspected dysfunctional cortical visual-vestibular integration in PPPD could be compensated by an adaptive increase of parieto-opercular FC with the caudal cerebellar vermis. However, there was no significant correlation with behavioral or perceptual parameters.

The larger FC of the patients’ visual motion sensitive MT/V5 and cerebellar Crus I might interfere with visual motion prediction. Area V5 is the key visual cortical region for motion processing. Dynamic causal modeling studies have shown enhanced connectivity between Crus I and V5 during visual attention, indicating that V5 becomes more sensitive to cerebellar Crus I input during attention-to-motion tasks^66^. At the same time, the modulatory influence of V5 on posterior parietal cortex activity was reduced, resulting in diminished suppression of vestibular bottom-up input. Accordingly, the abnormal visual attention observed in PPPD may alter the sensitivity of V5 to cerebellar prediction signals conveyed through enhanced FC with Crus I, particularly under conditions of increased uncertainty during motion perception^67^. Consistent with this interpretation, sensitivity of V5 to cerebellar input has been shown to increase when visuomotor adaptation to delayed visual feedback is required^68^.

We also identified a larger FC between OP3 and Crus II. OP3 is located within the secondary somatosensory cortex in the parietal operculum and has been shown to be activated during galvanic vestibular stimulation^69^ and stimulus-compatible egomotion^17^. Cerebellar Crus II is involved in spatial memory^70^ and contributes to the accurate temporal prediction of timing in voluntary movements^71^.

The larger FC between OP4 and the thalamus might reflect an increased reliance of spatial orientation of PPPD patients on vestibular and somatosensory rather than visual signal (reduced FC between Ig2 and lingual gyrus). OP4 is more closely integrated with areas responsible for basic sensorimotor processing and action control. It is heavily interconnected with the thalamus to integrate sensory, i.e., vestibular and somatosensory functions^72^.

### fALFF alteration in hippocampus

Fractional amplitude of low frequency fluctuations (fALFF) was larger in our patients’ right hippocampus. This has not been found in related studies yet. However, seed-based FC was found to be larger between the patients’ right hippocampus and the left frontal pole^12^, while it was reduced between the left hippocampus and the parietal operculum^16^. Impairments in spatial orientation, spatial memory, and navigation have long been linked to hippocampal dysfunction, and hippocampal atrophy may follow peripheral vestibular disorders^73, 74^. Recently, patients with PPPD despite normal vestibular function have been shown to exhibit deficits in allocentric spatial navigation^75^. Increased gaze scanning was interpreted as a compensatory strategy for spatial uncertainty. Accordingly, the larger connectivity of the right hippocampus may reflect a compensatory response to impaired spatial orientation. Further evidence for spatial orientation deficits in PPPD comes from behavioral 3D navigation tasks, in which impaired performance was attributed to a central suppression of vestibular input, resembling bilateral vestibulopathy, thereby interfering with the continuous updating of internal representations of body motion and spatial orientation relative to the environment^76^.

## Conclusion

The persistence of PPPD symptoms in the absence of peripheral sensory hypofunction suggests a central maladapative modification of the mechanisms involved in egomotion perception and postural control. This likely involves dysfunctional multisensory integration with abnormal intersensory weighting and aberrant precision weighting of sensory prediction errors. We provide evidence for abnormally stronger cerebro-cerebellar FC in PPPD, which may contribute to altered processing of sensory prediction errors leading to sensory-perceptual amplification, postural misperception, and a mismatch between predicted (perceived) and actual (recorded) egomotion. Specifically, we identified several cerebellar clusters functionally connected to cortical visual and vestibular regions known to be involved in egomotion and encoding of prediction errors. Concurrently, reduced visual-vestibular cortical FC may provide the error signals for this maladaptation. Whether the decrease of cortical visual-vestibular and the increase of cerebro-cerebellar FC are linked remains to be determined in future follow-up investigations.

## Author contributions

Conceptualization: HS, RS, CH, AS; Formal Analysis: HS, RS, AS, CH; Investigation: all authors; Methodology: HS, RS, CH, AS; Project Administration: CH, AS; Software: HS, AS; Supervision: CH, AS; Visualization: HS, RS, AS, CH; Writing – original draft: HS, CH, RS; Writing – review & editing: all authors.

## Data Availability

The data are available from the authors upon reasonable request.

## Acknowledgements

The author RS gratefully acknowledges support from the Detlef Zillikens Clinician Scientist Academy of Precision Health in Schleswig-Holstein (PHSH) and the Clinician Scientist Academy Lübeck.

Funding: This study was supported by a grant of the Deutsche Forschungsgemeinschaft (German Research foundation) to C.H. (HE 2689/6-1).

## Data availability

Neurological-clinical, behavioral (questionnaire, posturography, rating) and fMRI data are not publicly available to preserve individual’s privacy. The data are, however, available from the authors upon reasonable request. Analysis scripts are publicy available in a github repository: https://github.com/hannah-swe/fMRI_tools.

## Ethics approval

The study protocol was approved by the local Ethics Committee of the University of Lübeck (AZ 17–036, AZ 21–098) in accordance with the Declaration of Helsinki. Written informed consent was obtained from all participants.

## Abbreviations

PPPD: persistent postural-perceptual dizziness
HC: healthy controls
FC: functional connectivity
fALFF: fractional amplitude of low-frequency fluctuations
V5: Visual area 5
IPL: Inferior parietal lobule PFcm
OP1: Parietal operculum area OP1
OP3: Parietal operculum area OP3
OP4: Parietal operculum area OP4
Ig2: Insula Ig2
Crus I: Cerebellar Crus I
Crus II: Cerebellar Crus II
Lobule VI: Cerebellar lobule VI
Verm VIIIa: Vermis VIIIa
Verm IX: Vermis IX
SMG: Supramarginal gyrus
LingG: Lingual gyrus
Put: Putamen
Thal: Thalamus
Caud: Caudate nucleus
Hipp: Hippocampus
l/r: left/right

## Supplemental Materials

**Table S1.**
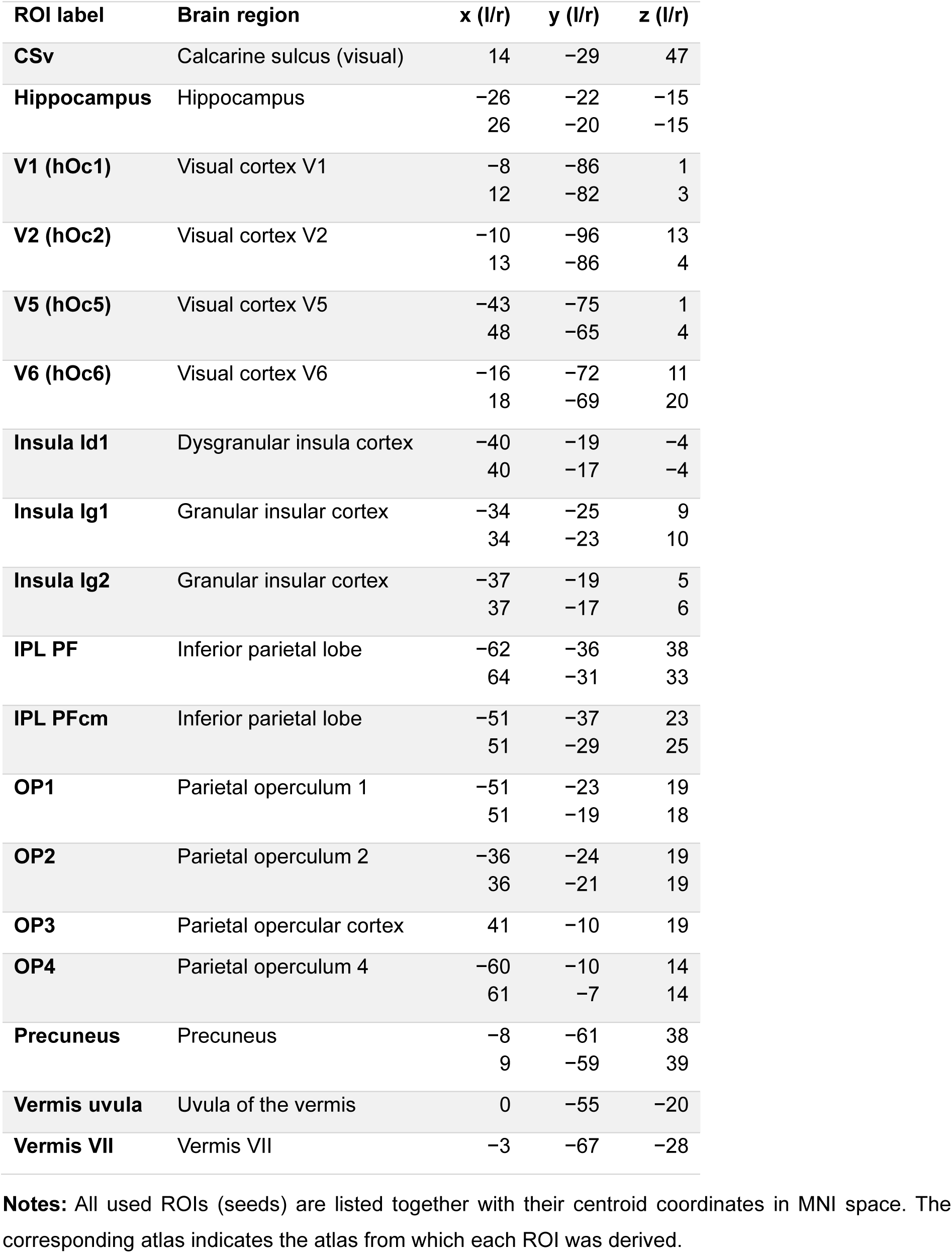
Regions of interest used for seed-based functional connectivity analyses.

| ROI label | Brain region | x (l/r) | y (l/r) | z (l/r) |
| --- | --- | --- | --- | --- |
| **CSv** | Calcarine sulcus (visual) | 14 | -29 | 47 |
| **Hippocampus** | Hippocampus | -26 | -22 | -15 |
|  |  | 26 | -20 | -15 |
| **V1 (hOc1)** | Visual cortex V1 | -8 | -86 | 1 |
|  |  | 12 | -82 | 3 |
| **V2 (hOc2)** | Visual cortex V2 | -10 | -96 | 13 |
|  |  | 13 | -86 | 4 |
| **V5 (hOc5)** | Visual cortex V5 | -43 | -75 | 1 |
|  |  | 48 | -65 | 4 |
| **V6 (hOc6)** | Visual cortex V6 | -16 | -72 | 11 |
|  |  | 18 | -69 | 20 |
| **Insula Id1** | Dysgranular insula cortex | -40 | -19 | -4 |
|  |  | 40 | -17 | -4 |
| **Insula Ig1** | Granular insular cortex | -34 | -25 | 9 |
|  |  | 34 | -23 | 10 |
| **Insula Ig2** | Granular insular cortex | -37 | -19 | 5 |
|  |  | 37 | -17 | 6 |
| **IPL PF** | Inferior parietal lobe | -62 | -36 | 38 |
|  |  | 64 | -31 | 33 |
| **IPL PFcm** | Inferior parietal lobe | -51 | -37 | 23 |
|  |  | 51 | -29 | 25 |
| **OP1** | Parietal operculum 1 | -51 | -23 | 19 |
|  |  | 51 | -19 | 18 |
| **OP2** | Parietal operculum 2 | -36 | -24 | 19 |
|  |  | 36 | -21 | 19 |
| **OP3** | Parietal opercular cortex | 41 | -10 | 19 |
| **OP4** | Parietal operculum 4 | -60 | -10 | 14 |
|  |  | 61 | -7 | 14 |
| **Precuneus** | Precuneus | -8 | -61 | 38 |
|  |  | 9 | -59 | 39 |
| **Vermis uvula** | Uvula of the vermis | 0 | -55 | -20 |
| **Vermis VII** | Vermis VII | -3 | -67 | -28 |

**Notes:** All used ROIs (seeds) are listed together with their centroid coordinates in MNI space. The corresponding atlas indicates the atlas from which each ROI was derived.

**Table S2.**
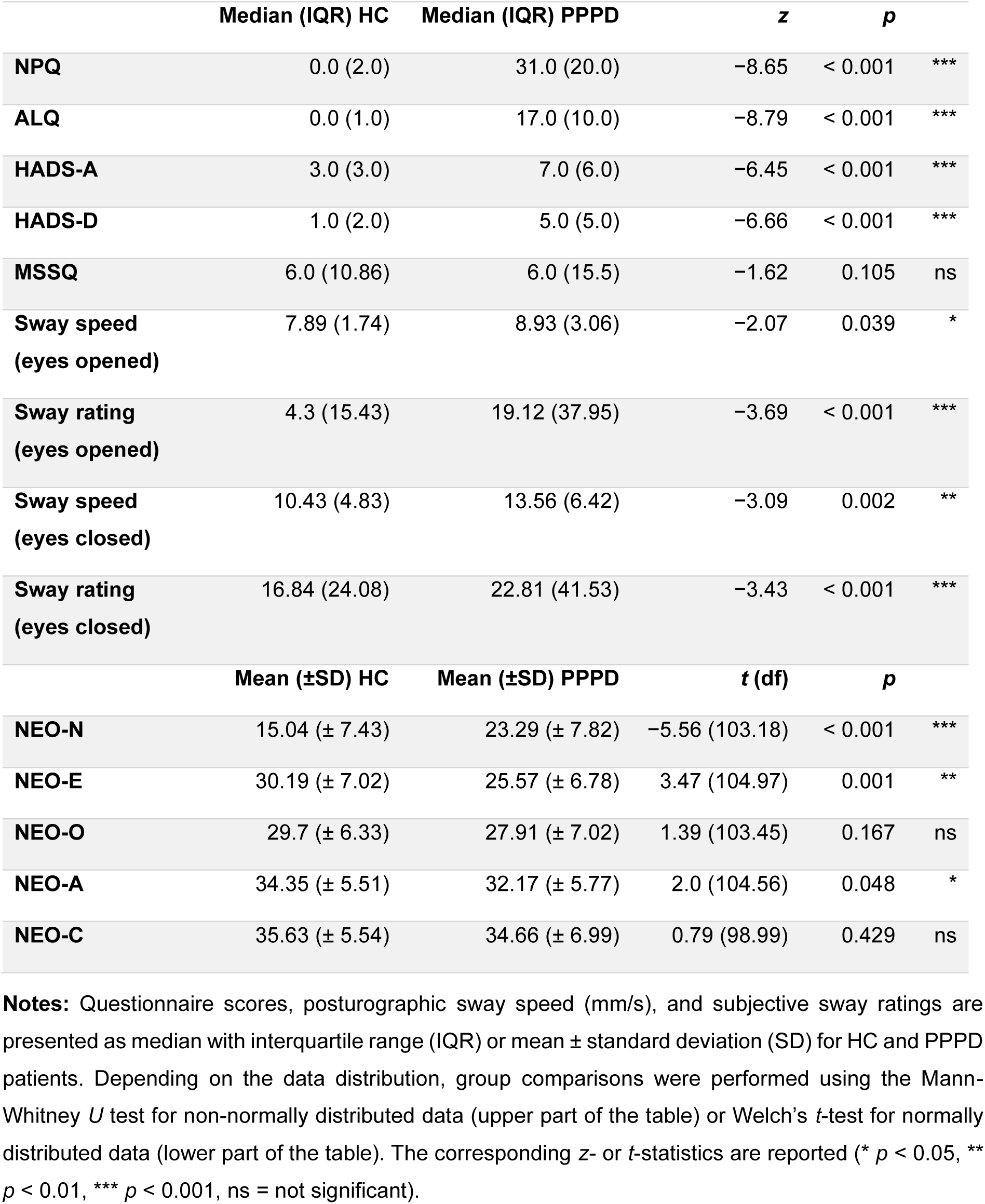
Questionnaire scores and posturography measures from HC and PPPD patients.

|  | Median (IQR) HC | Median (IQR) PPPD | *z* | *p* |  |
| --- | --- | --- | --- | --- | --- |
| **NPQ** | 0.0 (2.0) | 31.0 (20.0) | -8.65 | < 0.001 | *** |
| **ALQ** | 0.0 (1.0) | 17.0 (10.0) | -8.79 | < 0.001 | *** |
| **HADS-A** | 3.0 (3.0) | 7.0 (6.0) | -6.45 | < 0.001 | *** |
| **HADS-D** | 1.0 (2.0) | 5.0 (5.0) | -6.66 | < 0.001 | *** |
| **MSSQ** | 6.0 (10.86) | 6.0 (15.5) | -1.62 | 0.105 | ns |
| **Sway speed**<br>**(eyes opened)** | 7.89 (1.74) | 8.93 (3.06) | -2.07 | 0.039 | * |
| **Sway rating**<br>**(eyes opened)** | 4.3 (15.43) | 19.12 (37.95) | -3.69 | < 0.001 | *** |
| **Sway speed**<br>**(eyes closed)** | 10.43 (4.83) | 13.56 (6.42) | -3.09 | 0.002 | ** |
| **Sway rating**<br>**(eyes closed)** | 16.84 (24.08) | 22.81 (41.53) | -3.43 | < 0.001 | *** |
| | Mean (±SD) HC | Mean (±SD) PPPD | *t* (df) | *p* | |
| **NEO-N** | 15.04 (± 7.43) | 23.29 (± 7.82) | -5.56 (103.18) | < 0.001 | *** |
| **NEO-E** | 30.19 (± 7.02) | 25.57 (± 6.78) | 3.47 (104.97) | 0.001 | ** |
| **NEO-O** | 29.7 (± 6.33) | 27.91 (± 7.02) | 1.39 (103.45) | 0.167 | ns |
| **NEO-A** | 34.35 (± 5.51) | 32.17 (± 5.77) | 2.0 (104.56) | 0.048 | * |
| **NEO-C** | 35.63 (± 5.54) | 34.66 (± 6.99) | 0.79 (98.99) | 0.429 | ns |

**Notes:** Questionnaire scores, posturographic sway speed (mm/s), and subjective sway ratings are presented as median with interquartile range (IQR) or mean ± standard deviation (SD) for HC and PPPD patients. Depending on the data distribution, group comparisons were performed using the Mann-Whitney *U* test for non-normally distributed data (upper part of the table) or Welch's *t*-test for normally distributed data (lower part of the table). The corresponding *z*- or *t*-statistics are reported (\* *p* < 0.05, \*\* *p* < 0.01, \*\*\* *p* < 0.001, ns = not significant).

**Table S3.**
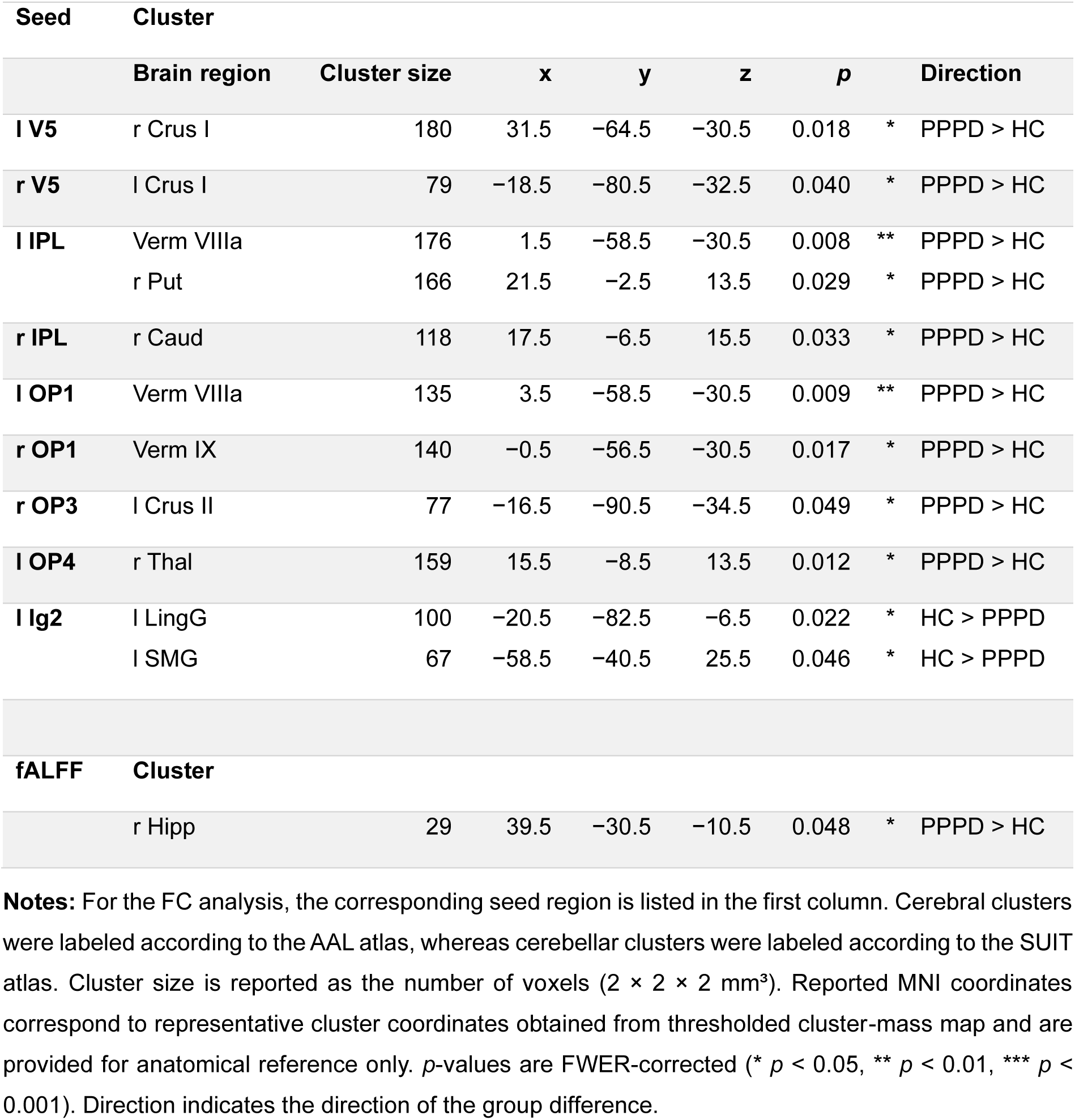
Significant clusters showing group differences in seed-based functional connectivity and fALFF analyses.

**Table S4.** Percentage of overlap of significant cerebellar clusters with SUIT atlas regions.

| Seed | x | y | z | SUIT label | Percentual overlap |
| --- | --- | --- | --- | --- | --- |
| **l V5** | 31.5 | -64.5 | -30.5 | Right Crus I | 65.5 % |
|  |  |  |  | Right lobule VI | 33.3 % |
| **r V5** | -18.5 | -80.5 | -32.5 | Left Crus I | 55.2 % |
|  |  |  |  | Left Crus II | 44.8 % |
| **l IPL** | 1.5 | -58.5 | -30.5 | Vermis VIIIa | 25.1 % |
|  |  |  |  | Vermis IX | 16.7 % |
|  |  |  |  | Right interposed nucleus | 8.7 % |
|  |  |  |  | Vermis VI | 7.5 % |
|  |  |  |  | Right lobule V | 6.7 % |
| **l OP1** | 3.5 | -58.5 | -30.5 | Vermis VIIIa | 22.2 % |
|  |  |  |  | Vermis IX | 16.6 % |
|  |  |  |  | Right interposed nucleus | 13.4 % |
|  |  |  |  | Right lobule V | 8.6 % |
|  |  |  |  | Vermis VIIIb | 8.2 % |
|  |  |  |  | Vermis VI | 7.4 % |
| **r OP1** | -0.5 | -56.5 | -30.5 | Vermis IX | 21.1 % |
|  |  |  |  | Vermis VIIIa | 18.4 % |
|  |  |  |  | Left lobule IX | 13.9 % |
|  |  |  |  | Vermis VI | 11.0 % |
|  |  |  |  | Right lobule V | 5.5 % |
| **r OP3** | -16.5 | -90.5 | -34.5 | Left Crus II | 74.7 % |

**Notes:** For each significant cerebellar cluster, the percentage of overlap with cerebellar regions defined by the SUI atlas is reported. The first column indicates the corresponding seed region. Coordinates are given in MNI space. Cluster labels were assigned to the cerebellar region with the largest overlap. Only overlaps greater than 5 % are reported.

**Figure S1.**
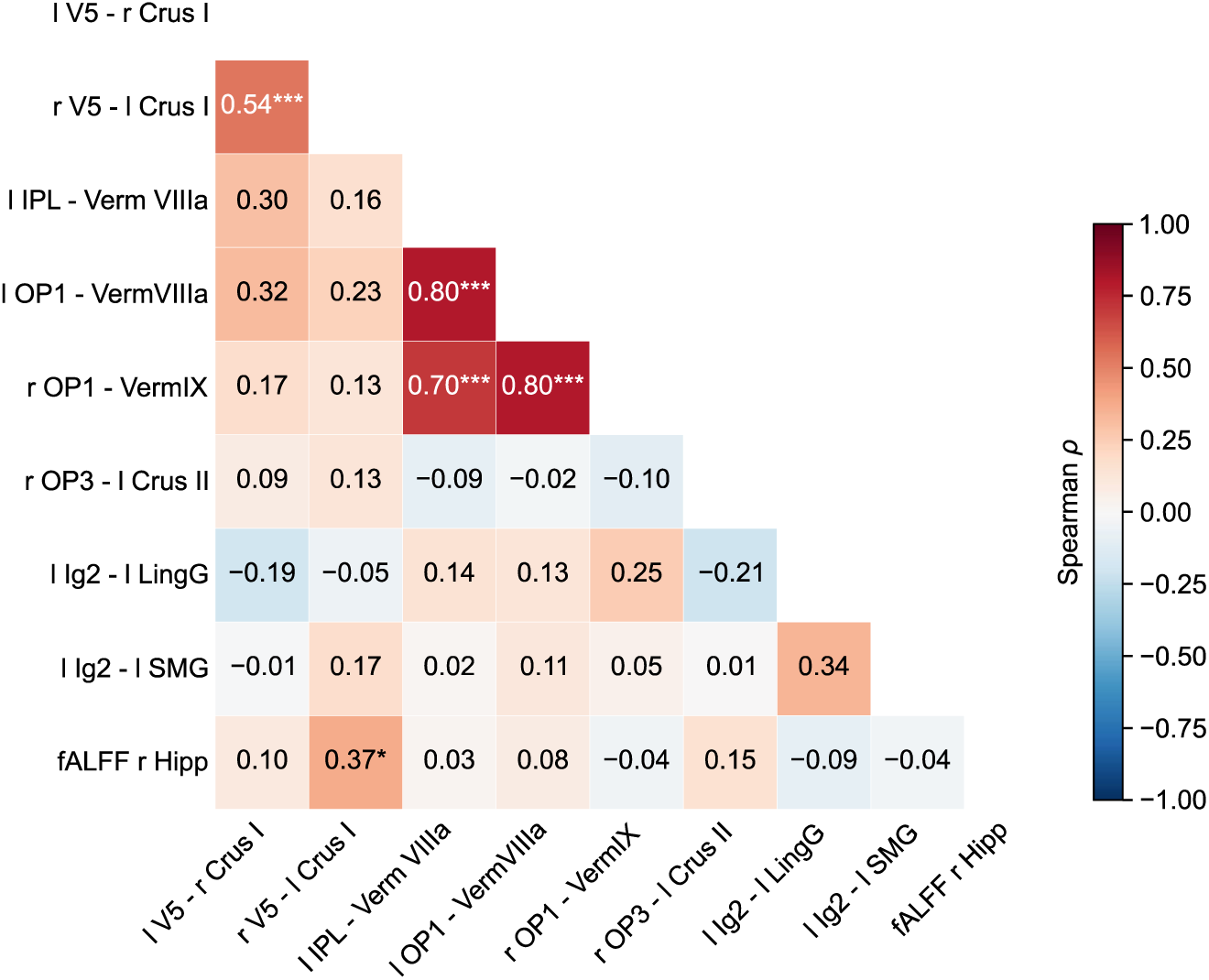
Correlation matrix of functional connectivity and fALFF measures in PPPD patients. The full correlation matrix includes all pairwise correlations between seed-based FC and fALFF values extracted from clusters showing significant group differences between PPPD patients and HC. Only the lower triangular part of the correlation matrix is shown. Each cell displays Spearman’s correlation coefficient (*ρ*). Asterisks indicate statistically significant correlations (* *p* < 0.05, ** *p* < 0.01, *** *p* < 0.001). *p*-values were corrected for multiple comparisons using FDR.

